# Molecular Characterization and Clinical Profile of Dengue Virus Serotypes in NS1-Positive Patients: A Cross-sectional Study from Rajkot, Gujarat, India

**DOI:** 10.64898/2026.03.16.26348474

**Authors:** Abhishek Padhi, Jignasa H. Bera, Bhoomika Rajyaguru, Jagruti Chauhan, Disha Rank, Ishita Modasiya, Sneh Bhalani, Ashwini Agarwal

**Affiliations:** Department of Microbiology, All India Institute of medical Sciences (AIIMS), Rajkot, Gujarat, India; Viral Research and Diagnostic Laboratory (VRDL), Department of Microbiology, (AIIMS), Rajkot, Gujarat, India

**Keywords:** Dengue virus, serotyping, RT-PCR, NS1 antigen, epidemiology, Gujarat, India

## Abstract

**Background:** Dengue virus infection remains a significant public health concern in India, with changing serotype dynamics influencing disease epidemiology. Understanding local serotype distribution and clinical characteristics is crucial for effective disease management and surveillance.

**Objectives:** To determine the prevalence of dengue virus serotypes and analyze their clinical characteristics among NS1-positive patients at a tertiary-care hospital in Gujarat, India.

**Methods:** A cross-sectional study was conducted on NS1-positive dengue patients admitted to AIIMS Rajkot from September 2023 to November 2024. Real-time reverse transcription polymerase chain reaction (RT-PCR) was performed for serotype identification. Clinical and demographic data were collected and analyzed.

**Results:** NS1-positive patients (70) were confirmed by RT-PCR. DENV-2 was the predominant serotype (53 cases, 75.7%), followed by DENV-1 and DENV-3 (7 cases each, 10.0%), and DENV-4 (2 cases, 2.9%). One co-infection case (DENV-2 + DENV-3) (1.4%) was identified. The mean age was 27.7 ± 14.4 years, with male predominance (58.6%). Young adults (19-35 years) were most affected (45.7%), followed by pediatric patients ≤18 years (32.9%). Severe dengue occurred in only one case (1.4%), while hospitalization was required in 25 cases (35.7%). All patients presented with fever, chills, headache (50%), rashes (56%), and malaise (56%), being the most common associated symptoms.

**Conclusions:** DENV-2 showed clear predominance in the Rajkot region during the study period, with low rates of severe disease. The significant pediatric and young adult involvement highlights the need for targeted prevention strategies. These findings contribute to the understanding of regional dengue epidemiology and support evidence-based surveillance and control measures.

## Introduction

Dengue virus (DENV) infection represents one of the most rapidly spreading mosquito-borne diseases globally, with an estimated 390 million infections annually [1]. In India, dengue has emerged as a major public health challenge, with all four serotypes (DENV-1 to DENV-4) circulating and causing periodic outbreaks [2,3]. The disease burden has increased substantially over the past decades, with changing epidemiological patterns and serotype dominance across different regions [4].

The clinical presentation of dengue ranges from asymptomatic infection to severe manifestations, including dengue hemorrhagic fever (DHF) and dengue shock syndrome (DSS) [5]. Serotype-specific differences in clinical manifestations have been documented, with DENV-2 infections often associated with more severe disease outcomes [6,7]. Understanding regional serotype distribution is crucial for predicting outbreak patterns, assessing disease severity risk, and planning appropriate public health responses [8].

Hyperendemicity is defined as the simultaneous circulation of all four dengue serotypes within a geographic region [5]. This epidemiological pattern significantly increases the risk of severe dengue through secondary infections due to antibody-dependent enhancement (ADE) and other immunopathological mechanisms [9,10]. Regions experiencing hyperendemic transmission require enhanced surveillance and clinical preparedness.

Gujarat, located in western India, has experienced several dengue outbreaks in recent years. The state reported varying seropositivity rates, with 4.7% in 2023 and 3.5% in 2024, indicating ongoing transmission [11]. However, comprehensive molecular characterization data from the Saurashtra region, particularly from tertiary care centers, remain limited.

The NS1 antigen-based rapid diagnostic tests have become the primary screening tool for early dengue diagnosis, with sensitivities ranging from 50-90% depending on the serotype and timing of testing [12,13]. Molecular confirmation through RT-PCR provides definitive diagnosis and enables serotype identification, which is essential for epidemiological surveillance [14].

Previous studies from India have shown varying serotype predominance across different states and time periods. While DENV-2 has been frequently reported as the dominant serotype in North and West India, regional variations exist [15,16]. In Gujarat specifically, limited recent data exist on molecular serotype characterization from healthcare facilities.

This study aims to address this knowledge gap by providing a comprehensive molecular and clinical characterization of dengue cases from AIIMS Rajkot, a major tertiary care center serving the Saurashtra region of Gujarat. The objectives were to: (1) determine the prevalence and distribution of dengue virus serotypes among NS1-positive patients, (2) analyze clinical and demographic characteristics associated with different serotypes, and (3) assess disease severity patterns in the study population.

## Materials and Methods

### Study Design and Setting

This cross-sectional observational study was performed at the All India Institute of Medical Sciences (AIIMS), Rajkot, Gujarat, India. This study was conducted by following the Strengthening the Reporting of Observational Studies in Epidemiology (STROBE) guidelines [15]. AIIMS, Rajkot, is a tertiary care hospital serving the Saurashtra region of Gujarat and receives patients from the Rajkot district and surrounding areas.

### Study Period and Population

The study included all NS1 antigen-positive dengue patients presenting to AIIMS Rajkot from September 2023 to November 2024. Patients of all ages with confirmed NS1 positivity were included. Exclusion criteria included patients with incomplete clinical data or those who did not consent for additional testing.

### Sample Collection and Processing

Venous blood samples (3-5 ml) were collected in plain tubes within 1-7 days of fever onset. NS1 antigen testing was performed using commercially available rapid diagnostic test and Monalisa ELISA kits as per the manufacturer’s protocol (Bio-Rad). Serum samples from NS1-positive patients were stored at -80°C until RT-PCR analysis. Figure 1 depicts the workflow of dengue serotyping of NS1-positive patients.

**Figure 1.**
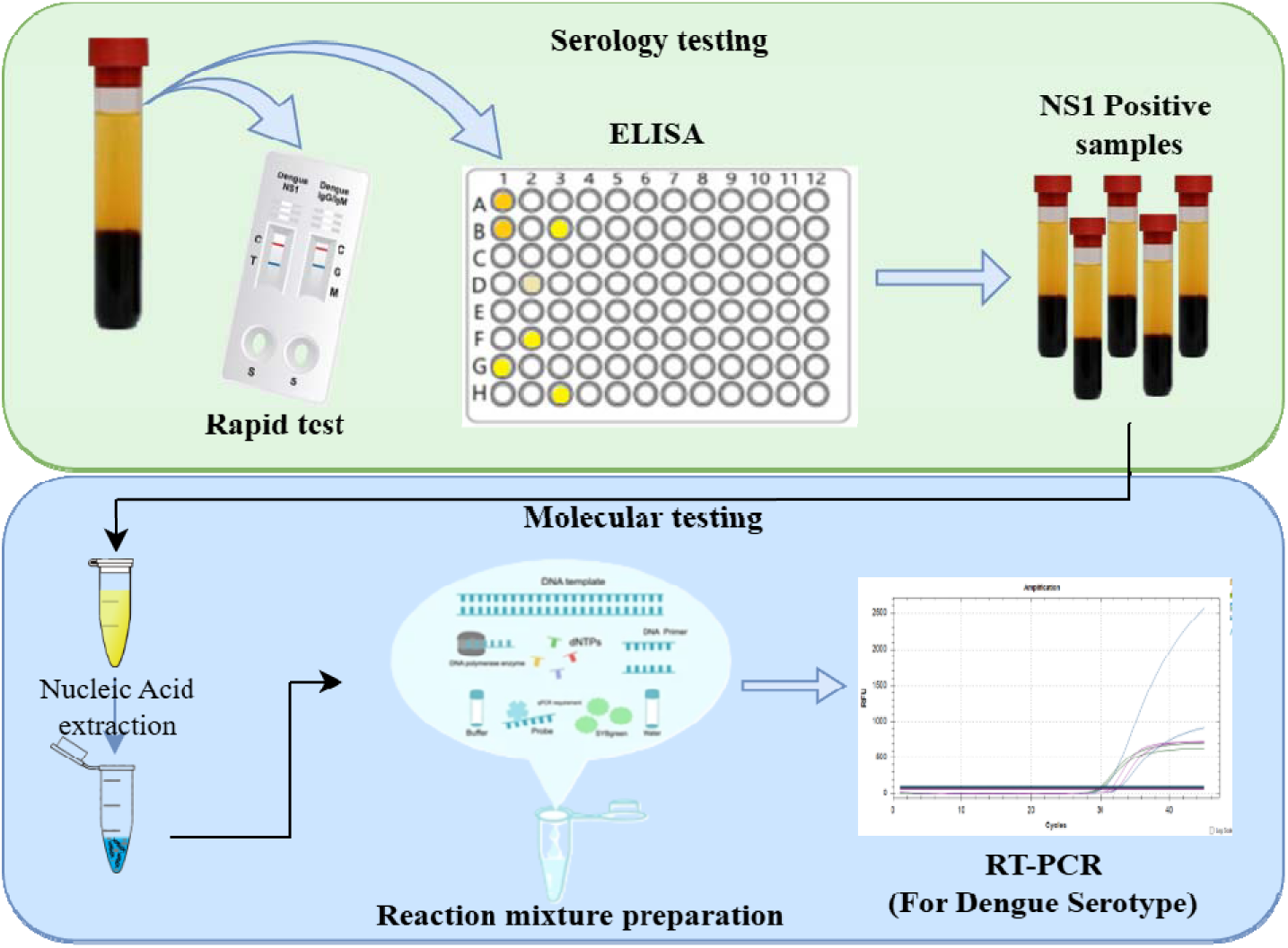
Representation of work flow forNS1 positive patients dengue serotyping

### Molecular Diagnosis and Serotyping

NS1 positive serum samples were sent to ICMR NIV for dengue virus detection and serotyping. Viral RNA extraction was performed using QIAamp RNA extraction kit (Qiagen, Germany) following the manufacturer’s instructions. Real-time RT-PCR for dengue virus detection and serotyping was conducted using validated primers and probes specific for DENV-1, DENV-2, DENV-3, and DENV-4 serotypes by RealStar® Dengue Type RT-PCR Kit 1.0 RUO (Altona Diagnostics GmbH, Germany). The RT-PCR protocol included reverse transcription at 55°C for 20 minutes, initial denaturation at 95°C for 2 minutes, followed by 45 cycles of denaturation at 95°C for 15 seconds, annealing at 55°C for 45 seconds, and extension at 72°C for 15 seconds as per the manufacturer’s instructions.

Cycle threshold (Ct) values ≤40 were considered positive for dengue virus RNA. Serotype determination was based on specific amplification curves and Ct values for each serotype-specific assay, based on the in-house validation studies for RUO kit.

### Clinical Data Collection

Standardized case record forms were used to collect demographic, clinical, and laboratory data including: 1) Demographics: age, gender, residence; 2) Clinical features: fever duration, symptoms (headache, myalgia, arthralgia, retro-orbital pain, rash, nausea, vomiting); 3) Disease severity: classification according to WHO 2009 guidelines 4) Laboratory parameters: platelet count, hematocrit, liver enzymes; 5) Hospitalization requirements and outcomes.

### Case Definitions

Cases were classified according to the WHO 2009 dengue case classification: **1) Dengue fever (DF):** Fever with two or more symptoms (headache, retro-orbital pain, myalgia, arthralgia, rash, hemorrhagic manifestations). **2)Dengue with warning signs:** DF with warning signs (abdominal pain, persistent vomiting, fluid accumulation, mucosal bleeding, lethargy, liver enlargement, increasing hematocrit with decreasing platelets). **3) Severe dengue:** Dengue with severe plasma leakage, severe bleeding, or severe organ involvement.

### Statistical Analysis

Data was analyzed using SPSS version 26.0 (IBM Corp., Armonk, NY). Descriptive statistics included frequencies, percentages, means ± standard deviation, and medians with ranges as appropriate. Chi-square test was used for categorical variables. Multivariable analysis was done to understand the factors associated with hospitalization. A p-value <0.05 was considered statistically significant.

### Ethical Considerations

The study was conducted in accordance with the Declaration of Helsinki and approved by the Institutional Ethics Committee of AIIMS Rajkot (IEC Approval No.: AIIMS/RAJKOT/5^th^ IEC/FB/36). Written informed consent was obtained from all adult participants or guardians for pediatric patients.

## Results

### Study Population Characteristics

A total of 70 NS1-positive patients were enrolled during the 14-month study period from September 2023 to November 2024. All 70 patients were confirmed positive by RT-PCR, indicating high concordance between NS1 antigen detection and molecular confirmation.

### Demographic Profile

The study population comprised 41 males (58.6%) and 29 females (41.4%), with a male-to-female ratio of 1.41:1. The mean age was 27.7 ± 14.4 years (median: 26 years, range: 1-80 years). Age group analysis revealed that young adults (19-35 years) were most affected (32 patients, 45.7%), followed by pediatric patients ≤18 years (23 patients, 32.9%). Adults aged 36-60 years comprised 13 patients (18.6%), while elderly patients >60 years represented only 2 cases (2.9%) (Table 1).

**Table 1.** Demographic and clinical characteristics of the confirmed dengue patients.

| Variable | Value |
| --- | --- |
| Total patients, n | 70 |
| Age (years), means ± SD | 27.7 ± 14.4 |
| Age (years), median (range) | 26 (1-80) |
| **Age Groups** |  |
| Pediatrics (≤ 18 years), n (%) | 23 (32.9) |
| Young adults (19-35 years), n (%) | 32 (45.7) |
| Adults (36-60 years), n (%) | 13 (18.6) |
| Elderly ($>$60years), n (%) | 2 (2.9) |
| **Gender Distribution** |  |
| Male, n (%) | 41 (58.6) |
| Female, n (%) | 29 (41.4) |
| Male: Female ratio | 1.41: 1 |
| **Clinical Outcomes** |  |
| Severe dengue, n (%) | 1 (1.4) |
| Hospitalization required, n (%) | 25 (35.7) |
| **Laboratory Findings** |  |
| NS1 antigen positive, n (%) | 70 (100.0) |
| RT-PCR positive, n (%) | 70 (100.0) |
| Ct value, mean ± SD | 31.7 ± 3.8 |
| Ct values, range | 22-39 |
| Duration of illness (days), mean ± SD | 3.9 ± 1.3 |

### Serotype Distribution and Molecular Characteristics

DENV-2 was the predominant serotype, accounting for 53 cases (75.7% of all confirmed cases). DENV-1 and DENV-3 each contributed 7 cases (10%), while DENV-4 was identified in 2 cases (2.9%). One case (1.4%) showed co-infection with DENV-2 and DENV-3 (Table 2). The RT-PCR Ct values ranged from 22 to 39 (mean: 31.7 ± 3.8), indicating varying viral loads among patients. The presence of all four serotypes in the Rajkot region indicates hyperendemic transmission.

**Table 2.** Serotype distribution and clinical outcomes among the dengue positive patients.

| Serotype | n (%) | Non-severe | Severe |
| --- | --- | --- | --- |
| DENV-2 | 53 (75.7) | 52 | 1 |
| DENV-1 | 7 (10.0) | 7 | 00 |
| DENV-3 | 7 (10.0) | 7 | 00 |
| DENV-4 | 2 (2.9) | 2 | 00 |
| DENV-2+ DENV-3 Co-infection | 1 (1.4) | 1 | 00 |
| **Total** | **70 (100.0)** | **69** | **1** |

### Clinical Manifestations

All patients presented with fever, which was the universal presenting symptom. The mean duration of illness at the time of sample collection was 3.9 ± 1.3 days (range: 2-7 days). The most common associated symptoms are observed in patients, as shown in Figure 2. This indicates that fever and chills were common in all patients. Where headache, malaise, and rashes were the second most common symptoms observed in patients.

**Figure 2.**
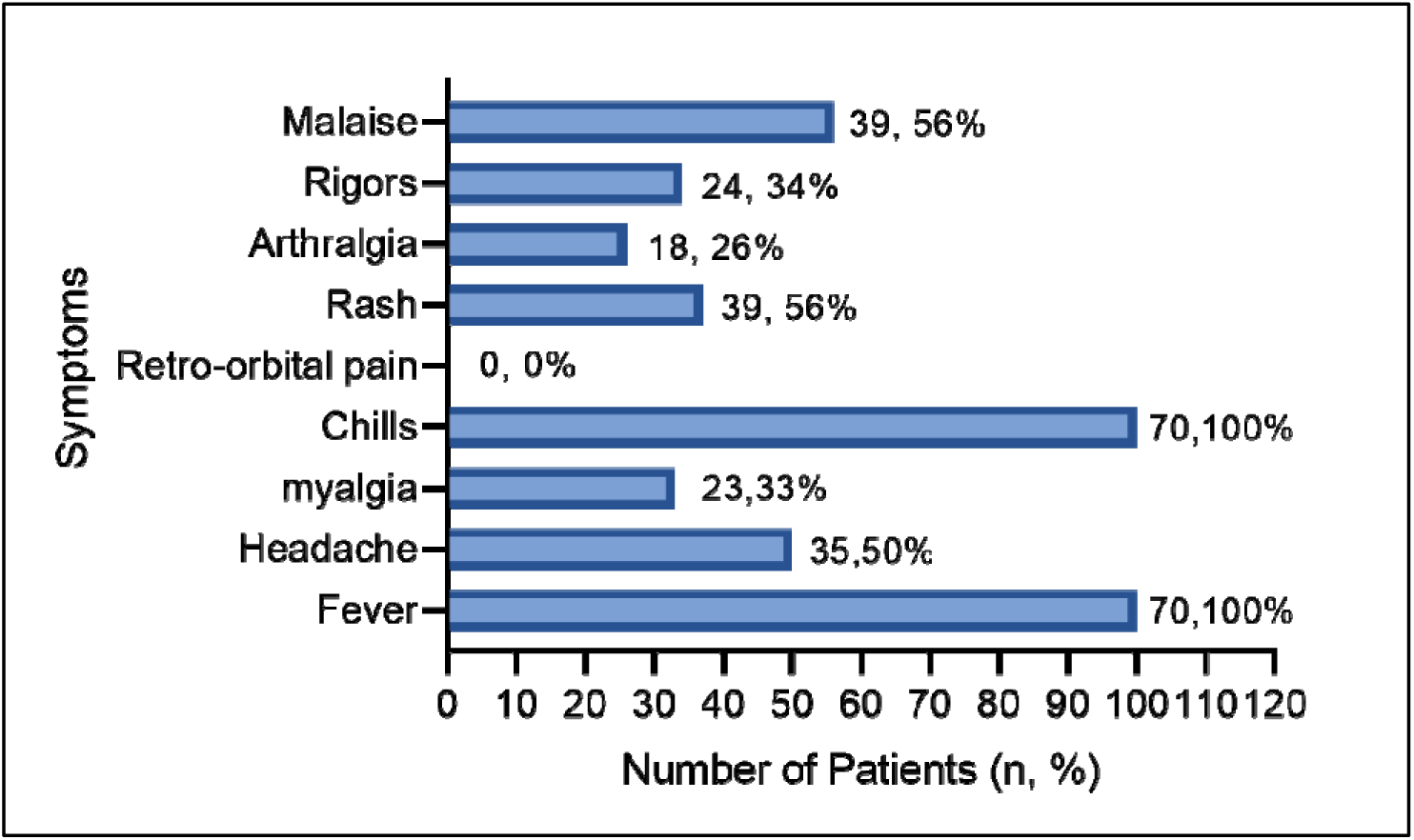
Frequency of symptoms observed among dengue patients (data are presented as number, %)

### Disease Severity and Clinical Outcomes

The majority of patients (69 cases, 98.6%) had uncomplicated dengue fever. Only one patient (1.4 %) developed severe dengue, presenting with warning signs including severe abdominal pain and plasma leakage. No cases of dengue shock syndrome or dengue hemorrhagic fever were observed during the study period.

Hospitalization was required for 25 patients (35.7%), while 45 patients (64.3%) were managed on an outpatient basis. The decision for hospitalization was primarily based on clinical assessment, platelet counts, and presence of warning signs rather than serotype-specific factors.

### Hematological Parameters

Thrombocytopenia (platelet count <1,50,000/μL) was observed in 47 patients (67.1%). The platelet counts ranged from 22,000 to 2,52,000/μL, with a median count of 1,25,000/μL. Severe thrombocytopenia (<50,000/μL) was noted in 14 patients (20.0%), but bleeding manifestations were rare, occurring in only 3 patients (4.3%). 13 (92.86%) out of 14 patients were hospitalized, and only one (7.14%) was a non-hospitalized patient.

### Temporal Distribution

The cases were distributed throughout the study period, with peak incidence observed during post-monsoon months (September-November) as shown in Figure 3. This pattern aligns with typical dengue seasonality in the region, correlating with favorable mosquito breeding conditions.

**Figure 3.**
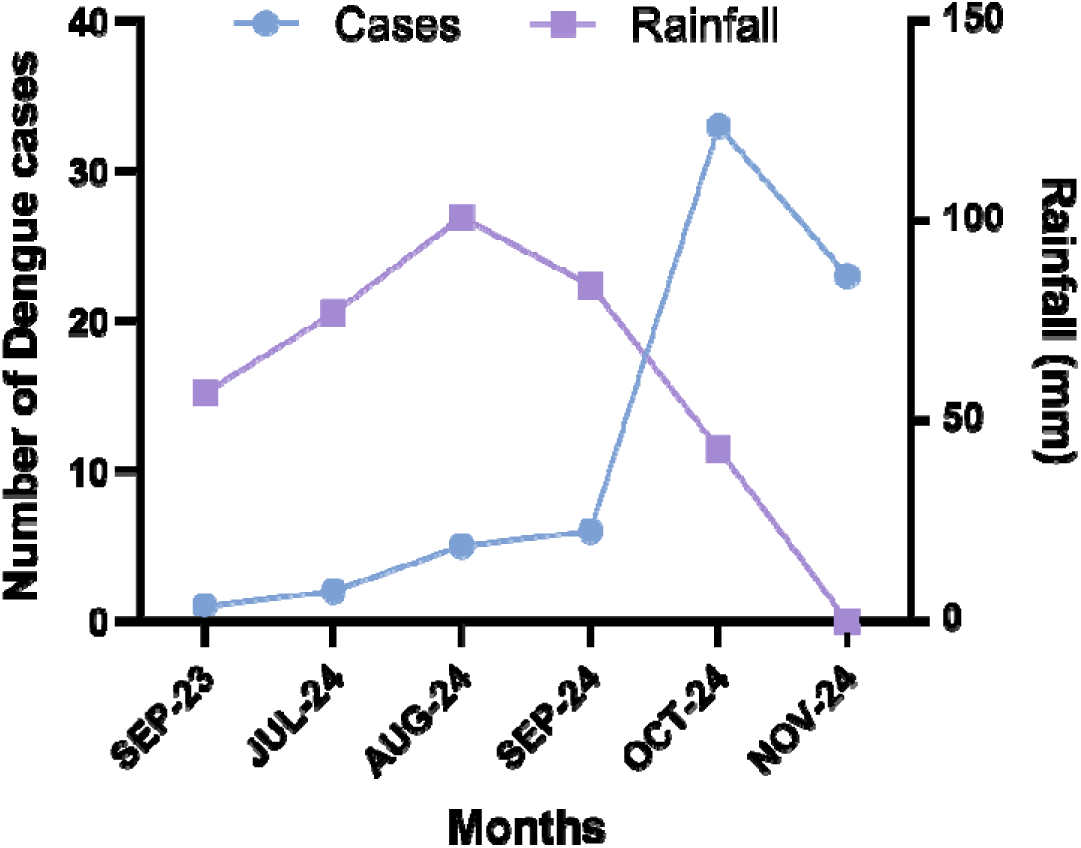
Distribution of dengue cases and rainfall pattern

### Age-Specific and Gender-Specific Patterns

Male patients showed a higher tendency for hospitalization compared to females (52.0% vs 48.0%), though this difference was not statistically significant (p=0.406). Adult patients (36.2%) had a slightly higher hospitalization rate compared to pediatric patients (34.8%), but this difference was not statistically significant (0.91).

### Multivariable analysis of factors associated with hospitalization

A multivariable logistic regression model was constructed, including age group, platelet count (per 10,000/ul increase), and Ct value. Hospitalization was independently associated with decreased platelet counts after adjustment (OR 0.84 per 10,000/ul increase; 95% CI 0.75-0.92; p= 0.0009). No significant correlation of hospitalization with age group (OR 1.02; 95% CI 0.32- 3.30; p=0.97) or Ct value (OR 0.99; 95% CI 0.86-1.14; p=0.91).

### Serotype-Specific Clinical Characteristics

No significant differences in clinical manifestations were observed between different serotypes, likely due to the small sample sizes for DENV-1, DENV-3, and DENV-4. However, the single severe dengue case was associated with DENV-2 infection, consistent with literature reports of DENV-2 being associated with more severe manifestations.

## Discussion

This study provides important insights into the molecular epidemiology and clinical characteristics of dengue virus infections in the Saurashtra region of Gujarat, India. The findings demonstrate a clear predominance of DENV-2 (77.1%) among NS1-positive patients, which is consistent with recent trends observed in various parts of India and globally [17][10].

### Serotype Distribution and Regional Patterns

The dominance of DENV-2 in our study aligns with reports from other Indian states, where DENV-2 has been identified as the predominant circulating serotype in recent years [18,19]. A recent systematic review of dengue seroprevalence in India reported that North and East India had higher dengue prevalence, with West India, including Gujarat, showing moderate prevalence rates [20]. Prior studies suggest that the predominance of DENV-2 in Gujarat is particularly significant as this serotype has been associated with more severe clinical outcomes and secondary infections [21]. The limited number of severe cases and the small number of other serotypes in our cohort study prevent a definitive conclusion regarding serotype-specific severity.

The detection of all four dengue serotypes in our study, albeit with varying frequencies, confirms the hyperendemic nature of dengue transmission in the region. The presence of co-infection (DENV-2 + DENV-3) in one patient highlights the circulation of multiple serotypes simultaneously, which has implications for disease severity and immune responses [22].

### Clinical Characteristics and Disease Severity

The clinical presentation in our study was consistent with typical dengue fever manifestations reported in the literature [23]. The universal presence of fever, high frequency of headache (50%), and myalgia (33%) matches the clinical description provided in national dengue management guidelines [24]. The relatively low incidence of severe dengue (1.4%) in our study is encouraging and may reflect early diagnosis, appropriate clinical management, or characteristics of the circulating viral strains.

The hospitalization rate of 35.7% in our study is comparable to other Indian studies, where hospitalization rates typically range from 30-50% depending on the healthcare setting and patient selection criteria [16]. The decision for hospitalization was primarily driven by clinical assessment and laboratory parameters rather than serotype-specific factors, which is appropriate given the current management protocols.

### Age and Gender Distribution

The predominance of young adults (19-35 years, 45.7%) and significant pediatric involvement (32.9%) in our study reflects the typical age distribution of dengue cases in endemic areas [25]. This pattern suggests ongoing transmission with periodic exposure of susceptible populations. The higher male-to-female ratio (1.41:1) observed in our study is consistent with many dengue studies from India, possibly reflecting occupational exposure patterns or healthcare-seeking behavior differences [26].

The substantial pediatric involvement (32.9%) is particularly noteworthy from a public health perspective, as children may have more severe manifestations and require different clinical management approaches [27]. This finding supports the need for enhanced vector control measures in residential areas and schools.

### Laboratory Findings and Molecular Characteristics

The high concordance (100%) between NS1 antigen positivity and RT-PCR confirmation validates the reliability of NS1-based screening in our setting. This finding is consistent with recent studies showing good agreement between NS1 rapid tests and RT-PCR, particularly when testing is performed within the first week of illness [28,29].

The mean Ct value of 31.7 ± 3.8 indicates moderate to high viral loads in most patients, which is expected given that samples were collected within 2-7 days of illness onset. The range of Ct values (22-39) suggests varying viral loads, which may reflect differences in the timing of sample collection relative to viremia peaks [30].

### Implications for Public Health and Clinical Practice

The predominance of DENV-2 has several important implications. First, it suggests the potential for more severe secondary infections if other serotypes become prevalent in the future, as DENV-2 antibodies may enhance infection severity through antibody-dependent enhancement (ADE) [31]. Second, it provides valuable information for dengue vaccine considerations, as vaccine efficacy may vary by serotype [32].

The low rate of severe dengue in our study, despite DENV-2 predominance, suggests that other factors such as host immunity, viral genotype, or early healthcare access may be protective. However, continued surveillance is essential as serotype dynamics can change rapidly [33].

### Comparison with Regional and National Data

Our findings are consistent with recent reports from Gujarat showing a decrease in dengue seropositivity rates from 4.7% in 2023 to 3.5% in 2024 [11]. However, the absolute number of cases in our study represents confirmed cases from a single tertiary center and may not reflect the true community burden of infection.

Compared to other Indian regions, the serotype distribution in Gujarat shows similarities with North and West India, where DENV-2 has been predominant in recent years [34]. This contrasts with some South Indian states where DENV-1 and DENV-3 have been more prevalent [35].

### Strengths and Limitations

The major strengths of this study include the use of molecular confirmation for all cases, comprehensive clinical data collection, and inclusion of all age groups. The study provides valuable baseline data for dengue serotype surveillance in Gujarat.

However, several limitations should be acknowledged. The study was conducted at a single tertiary care center, which may introduce selection bias toward more severe cases requiring hospital care. The sample size for individual serotypes other than DENV-2 was small, limiting statistical power for serotype-specific comparisons. Additionally, the study did not include serological testing for primary versus secondary infections, which would have provided additional insights into disease pathogenesis.

### Future Research Directions

Future studies should include multi-center surveillance across different healthcare levels in Gujarat to provide a more comprehensive picture of dengue epidemiology. Serological studies to determine primary versus secondary infection status would be valuable for understanding disease severity patterns. Genetic characterization of circulating viruses would provide insights into viral evolution and potential associations with clinical outcomes.

Long-term surveillance studies tracking serotype dynamics over multiple years would help predict outbreak patterns and inform public health preparedness. Additionally, studies investigating vector characteristics and entomological indices in the region would complement clinical surveillance data.

## Conclusions

This study demonstrates the clear predominance of DENV-2 among dengue cases in the Rajkot region of Gujarat, India, during 2023-2024. The low incidence of severe disease, despite DENV-2 dominance, is encouraging but requires continued monitoring. The significant involvement of pediatric and young adult populations highlights the need for targeted prevention strategies and enhanced vector control measures.

The high concordance between NS1 antigen testing and RT-PCR confirms the reliability of NS1-based screening for early dengue diagnosis in this setting. These findings contribute valuable data to the understanding of dengue epidemiology in West India and support evidence-based surveillance and control strategies.

Continued molecular surveillance is essential for tracking serotype dynamics, detecting emerging strains, and informing public health responses. The establishment of routine serotyping capabilities at regional centers would enhance dengue surveillance and support national disease control efforts.

## Data Availability

All data produced in the present work are contained in the manuscript

## Acknowledgments

We sincerely thank the Indian Council of Medical Research- National Institute of Virology (ICMR-NIV), Pune, for conducting the dengue serotyping. We sincerely acknowledge the support provided through the Indian Council of Medical Research (ICMR) and the Department of Health Research (DHR), Ministry of Health and Family Welfare, Government of India, under the VRDL scheme. We are also grateful to the Viral Research and Diagnostic Laboratory (VRDL), AIIMS Rajkot, where the study was carried out. The authors acknowledge the valuable technical assistance provided by Ms. Nikita Rathod, Laboratory Technician, VRDL, AIIMS Rajkot.

## Author Contributions

AP and AA conceptualized and designed the experiments. JC, and DR performed the experiments. AP, JC, JHB, IM, SB and BR analyzed the data. AP, AA, and BR validated and visualized data. AP and JHB wrote the original manuscript draft. All authors review final draft and approved for submission.

